# Heterogeneity and Risk-of-Biases are No Longer the Issues to Conclude the Effectiveness of Baricitinib in Reducing COVID-19 Related Mortality: A Systematic Review and Meta-Analysis of Eight Randomised Controlled Trials

**DOI:** 10.1101/2022.11.07.22282055

**Authors:** Sivananthan Manoharan, Lee Ying Ying

## Abstract

**Background:** Due to high heterogeneity and risk of bias (RoB) in previously published meta-analysis, a concrete conclusion on the efficacy of baricitinib in reducing mortality in COVID-19 patients was unable to form.

**Methods:** Search engines PubMed/MEDLINE, ScienceDirect and other sources like preprints and reference lists were searched with appropriate keywords. The included evidence was graded with GRADEpro. The RoB, heterogeneity and meta-analysis were studied through RevMan 5.4.1 software. The heterogeneity was evaluated based on the generated *p*-value or I² test.

**Results:** Eight (8) RCTs were included in current analysis. Five studies had low RoB. Based on grading the evidence, the inclusion and exclusion of high RoB articles led to moderate and high certainty of evidence, respectively. Based on 8 RCTs (with high RoB), baricitinib statistically significantly reduced mortality where the risk ratio (RR) = 0.84 [95% CI: 0.76 to 0.92; *p* = 0.0002; I^2^ = 23%; *p* = 0.25]. The heterogeneity was insignificant but the RoB was high. We did subgroup analysis of low and high RoB articles and found out baricitinib statistically significantly reduced mortality with the RR = 0.68 [95% CI: 0.56 to 0.82; *p* < 0.0001; I^2^ = 0%; *p* = 0.85] and RR = 0.89 [95% CI: 0.80 to 0.99; *p* = 0.04; I^2^ = 0%; *p* = 0.43], respectively. The heterogeneity was 0% with insignificant *p*-values in both subgroup analyses. The percentage of mortality reduction was 31.31% and 7.79%, respectively whereas it was 13.95% in main group analysis.

**Conclusion:** With the presence of optimal sample size of 3944 from 5 low RoB studies which represents a minimum of 300 million population of people and with 0% of heterogeneity, the effectiveness of baricitinib in reducing the mortality in COVID-19 patients is concretely proven.

## 1. Introduction

As of 06 November 2022, approximately 637 million of total COVID-19 cases have been reported worldwide which resulted in more than 6.6 million deaths [1]. The emergence of SARS-CoV-2 variants like Alpha, Beta, Gamma, Delta and Omicron further challenges the healthcare systems. The continuous efforts are being made by the scientists to discover new therapies for global citizens who are infected with SARS-CoV-2 virus. One of the strategies was to repurpose an anti-rheumatoid arthritis drug, baricitinib for the management of COVID-19 patients. Baricitinib, a Januse Kinase 1 (JAK 1) and 2 (JAK 2) inhibitor is the first immunomodulator which was found to reduce the mortality (death) in COVID-19 patients. Baricitinib was revealed to decrease various cytokines and biomarkers involved in COVID-19 pathophysiology. This drug is available in tablet form and due to its affordability, they are used in low and middle-income nations [2-3]. A recent meta-analysis (MA) based on 4 RCTs reported that baricitinib statistically significantly reduced mortality in COVID-19 patients [4]. In this MA, the authors had included RECOVERY study related data where RECOVERY study was an open label study which subject to high risk of bias (RoB). Furthermore, based on the MA analysis for mortality, the heterogeneity (I^2^) was significant with I^2^ = 65% and *p*-value = 0.04. When the RoB and heterogeneity are high/significant, the data need to be interpreted carefully. With the availability of latest ACTT-4 and several more RCTs data related to mortality, the MA needs to be updated in order to guide the clinicians with latest information related to baricitinib for the management of COVID-19 disease. In current work, the authors updated the systematic review (SR) and MA with the latest mortality data from 4 RCTs with the aim to derive a clear-cut conclusion for mortality.

## 2. Methodology

In the latest SR and MA, the Preferred Reporting Items for Systematic Reviews and Meta-Analyses (PRISMA) guidelines were followed in order to shape this review. The guidelines were followed accordingly. No advanced protocol associated to the latest SR and MA was made or registered.

### 2.1 Research questions

1. What is the statistical ability of baricitinib to reduce mortality/death in COVID-19 patients?
2. Does heterogeneity and RoB remain as obstacles to derive a clear-cut conclusion for mortality?

### 2.2 Search strategies, article eligibility criteria and data charting process

Two well-known databases, known as PubMed/MEDLINE, ScienceDirect and additional sources, such as preprints and reference list were explored systematically with keywords, specifically ‘Randomised controlled trials baricitinib COVID-19; Randomised controlled trials baricitinib SARS-CoV-2 virus; and Randomised controlled trials baricitinib pneumonia’. The search was done manually in preprint and reference list. The searched year was in between 2020 and 30 October 2022. The inclusion criteria were:

(i) Patients disease-ridden with SARS-CoV-2 virus;
(ii) Baricitinib was used for intervention purposes;
(iii) Compulsory existence of proper control/s;
(iv) Stringently only for RCT works;
(v) Clinical efficacy mentioned in the study results; and
(vi) Language restriction: Only English language articles.

The data charting process were undertaken entirely by two authors, independently. Both authors had no issue with any shortlisted articles and thus, not requiring the third person’s involvement to resolve the problem.

### 2.3 Risk of bias analysis (RoB), grading the evidence using GRADEpro and conduct of MA

The Cochrane RoB tool was used to determine the quality of RCTs [5]. Moreover, the authors conducted an additional step to grade the quality of the included articles with GRADEpro Guideline Development Tool software [6]. The MA was conducted via Review Manager 5.4.1 [5]. Dichotomous data type was selected. The data related to RCTs were pooled, and relative risk, confidence interval (CI) and Mantel-Haenszel statistical methods were employed. The heterogeneity was evaluated based on the generated *p*-value, the I² test. Heterogeneity was considered as significant when *p*<0.1 or I²>50% [7]. A fixed-effect or random-effect model was selected when the data were homogeneous or heterogeneous, respectively. The publication bias (PB) was not studied because only eight articles were available. A minimum of ten articles are needed to yield a good statistical related funnel plot to detect PB.

## 3. Results and discussions

Based on the literature search, 8 RCT articles were included in qualitative and quantitative analyses. Table 1 was adopted and modified from the authors’ own published MA manuscript in the Canadian Journal of Infectious Diseases and Medical Microbiology [8]. Based on table 1, baricitinib was divided into 2 doses known as 2 mg or 4 mg (depending on the health of the patients) and was given to the included COVID-19 participants for maximum of 14 days. The age of the participants in both groups was consistent in all studies except for 2 open labelled studies. But the age is consistent in all full RCTs with low RoB. This consistency is important in order to derive a good conclusion. Although some little variations in genetics may present in the overall study populations (about 0.1%) [9] but significant variation in age will be a major confounding factor for MA. By looking at the period of study, most studies started in mid to late 2020 and concluded in 2021 with two studies completed in March and May 2022. During this period, different variants of SARS-CoV-2 virus emerged and the authors of 8 RCTs did not mentioned which strain of viruses they were looking at. We think that it could be more than 1 strains present in most (or all) of the studies. This is because most of the SARS-CoV-2 strains emerged in 2020 with Omicron in 2021 [10]. Out of 8 RCTs, five of them categorised as low RoB. Three RCTs with open label type of study were revealed to have high RoB. This is because in open label study, both drug administrator and study participants are alert of drug and treatment given [11]. This will lead to bias to the outcome of the results.

**Table 1:**
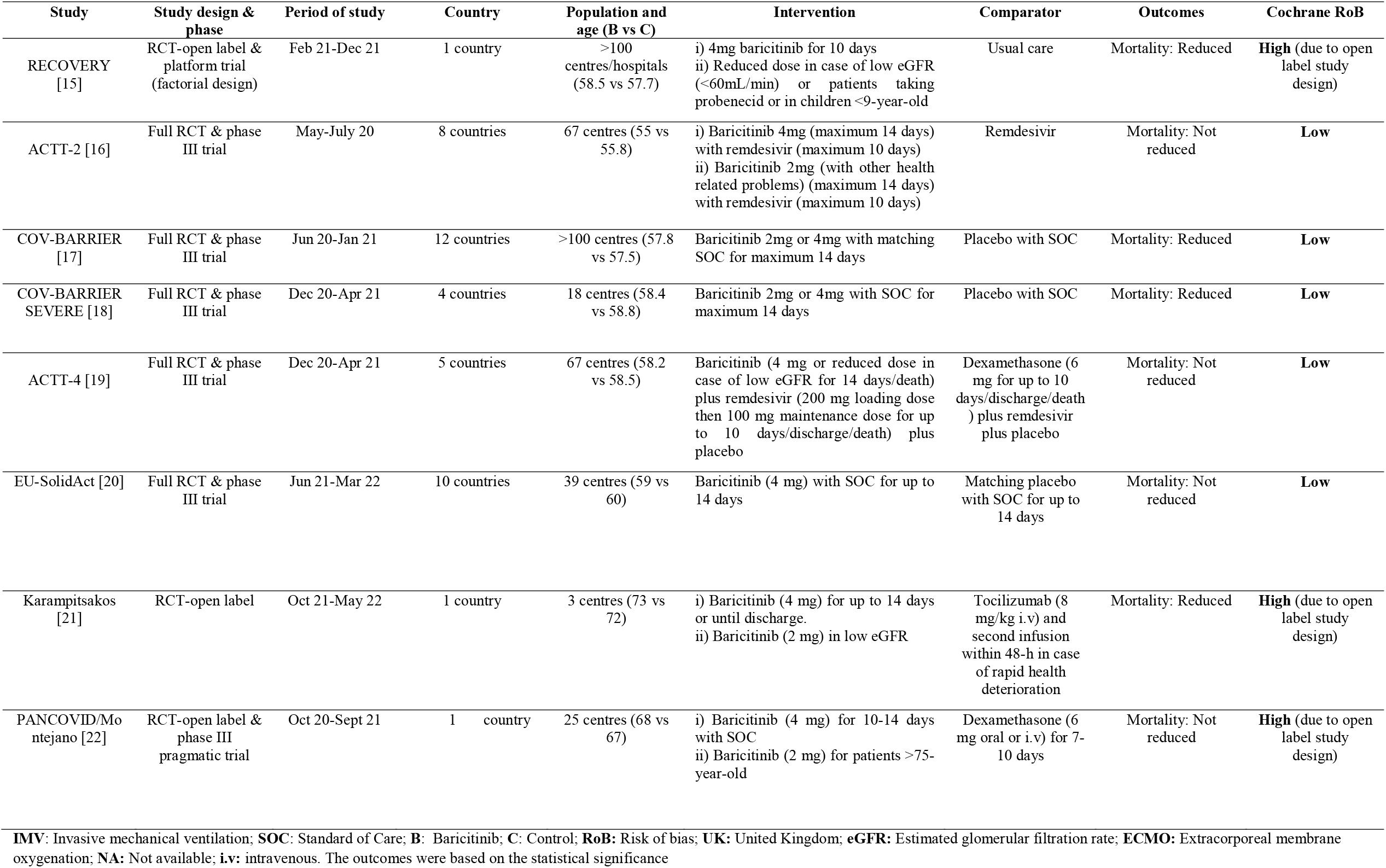
Characteristics of the included studies

Based on grading the evidences, the inclusion and exclusion of high RoB article led to moderate and high certainty of evidences, respectively (table 2). Inclusion of only high RoB articles produced moderate certainty of evidences. These 2 level of certainties in 3 different analyses gave different impressions of MA. In figure 1A where the moderate certainty present, the heterogeneity was low with I^2^ = 23% and *p* = 0.25. Fixed effect model was selected for the analysis. Treatment with baricitinib statistically significantly reduced mortality with the RR = 0.84 [95% CI: 0.76 to 0.92; *p* = 0.0002]. Although statistically significant, but since 3 high RoB articles present in the analysis, this analysis must be carefully interpreted. One of the ways to interpret is to look at table 1 and 2. The RoB and the certainty based on grading. When the RoB is high the certainty level is downgraded thus, leads to moderate quality/certainty of output due to performance bias in 3 included studies.

**Table 2:** Grading the evidence with GRADEpro Guideline Development Tool

| Certainty assessment |  |  |  |  |  |  | No of patients |  | Effect |  | Certainty | Importance |
| --- | --- | --- | --- | --- | --- | --- | --- | --- | --- | --- | --- | --- |
| No of studies | Study design | Risk of bias | Inconsistency | Indirectness | Imprecision | Other considerations | Baricitinib | Placebo | Relative (95% CI) | Absolute (95% CI) |  |  |
| Mortality 8 RCTs Mixed RoB |  |  |  |  |  |  |  |  |  |  |  |  |
| 8 | Randomised trials | Serious<sup>a</sup> | Not serious | Not serious | Not serious | None | 703/6403 (11.0%) | 817/6235 (13.1%) | RR 0.84 (0.76 to 0.92) | 21 fewer per 1,000 (from 31 fewer to 10 fewer) | ⊕⊕⊕⊟<br>Moderate | CRITICAL |
| Mortality 5 RCTs Low RoB |  |  |  |  |  |  |  |  |  |  |  |  |
| 5 | Randomised trials | Not serious | Not serious | Not serious | Not serious | None | 147/1985 (7.4%) | 214/1959 (10.9%) | RR 0.68 (0.56 to 0.82) | 35 fewer per 1,000 (from 48 fewer to 20 fewer) | ⊕⊕⊕⊕<br>High | CRITICAL |
| Mortality 3 RCTs High RoB |  |  |  |  |  |  |  |  |  |  |  |  |
| 3 | Randomised trials | Serious<sup>a</sup> | Not serious | Not serious | Not serious | None | 556/4418 (12.6%) | 603/4276 (14.1%) | RR 0.89 (0.80 to 0.99) | 16 fewer per 1,000 (from 28 fewer to 1 fewer) | ⊕⊕⊕⊟<br>Moderate | IMPORTANT |

CI: confidence interval; RR: risk ratio
Explanations
a. Open label study

**Figure 1:**
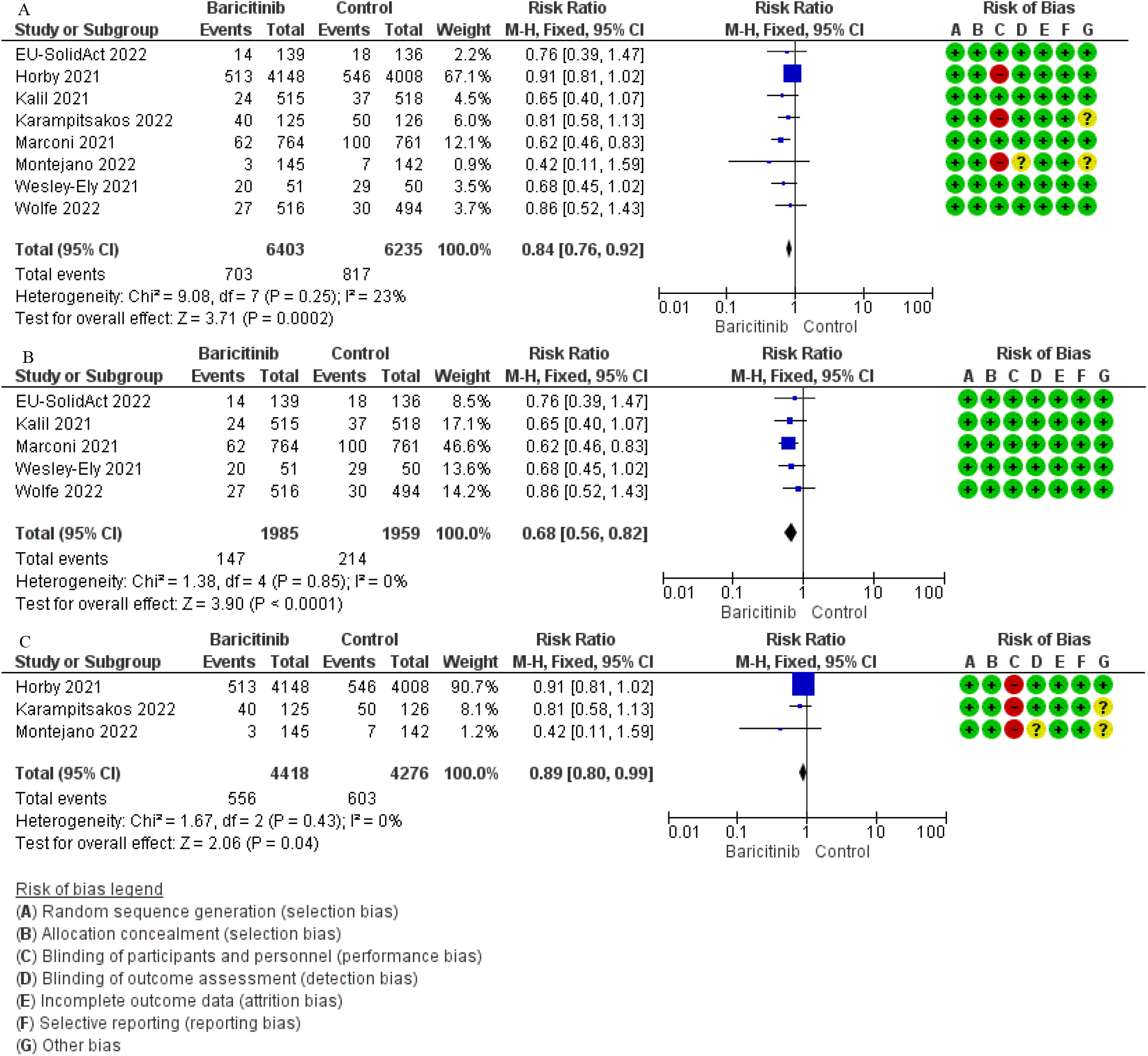
The outcome of baricitinib on mortality. (A) The MA of all included articles with and without RoB. (B) The MA of low RoB articles. (C) The MA of high RoB articles. On 29 October 2022, Karampitsakos [21] was contacted to clarify on whether the authors employed blind strategy for outcome assessment. The corresponding author replied yes, they did the blinding strategy to assess the outcome of the work. The similar email was sent to PANCOVID/Montejano [22] but no reply received.

Excluding 3 high RoB studies from the main analysis is another option to derive a solid conclusion for mortality which is based on 5 low RoB studies. Based on the calculated optimal information size (OIS) for figure 1B (data not shown), inclusion of 5 low RoB studies provided optimal information to derive conclusion. The total 3944 sample size in figure 2B yielded the required OIS. In fact, according to Eli Lilly, approximately 1 million patients received baricitinib for the management of COVID-19 disease in 15 countries [12]. With the margin of error of 2.5% and confidence = 95%, the sample size of 3944 represents a population of minimum 300 million [13]. In figure 1B, treatment with baricitinib statistically significantly reduced mortality with the RR = 0.68 [95% CI: 0.56 to 0.82; *p* < 0.0001; I^2^ = 0%; *p* = 0.85] with no heterogeneity was found. Based on figure 1B, with the presence of optimal sample size of 3944 from 5 low RoB studies which represents a minimum of 300 million population of people, the effectiveness of 2 mg or 4 mg baricitinib in reducing the mortality in COVID-19 patients was concretely proven. Despite large sample size, low heterogeneity, and significant reduction of mortality (*p*=0.04) by baricitinib in figure 1C but due to high performance bias and some other unclear biases, the effect of baricitinib in open-labelled RCTs need to be evaluated cautiously. Furthermore, RECOVERY study contributed the greatest number of participants plus events and the weightage of the study was 90.7% with the remaining weightage of just 8.3% is shared by another 2 small studies in figure 1C. Besides, based on table 1, karampitsakos [21] and PANCOVID/Montejano [22] produced different outcomes where the former mentioned baricitinib statistically reduced mortality while the latter mentioned the opposite. Based on the MA of these 2 studies, baricitinib statistically did not reduce mortality (data not shown) in COVID-19 patients where the RR = 0.76 [95% CI: 0.55 to 1.05; *p* = 0.1; I^2^ = 0%; *p* = 0.35]. With the margin of error of 2.5% and confidence = 95%, the total sample size of 538 in both studies represented just 800-1000 actual population [13]. Based on our observation, with the weightage of almost 91%, the produced result in figure 1C is mostly belongs to 1 study which is RECOVERY and the MA output should not be considered or used for any clinical interpretation by clinicians and/or by policy-makers. In conclusion, the availability of additional 4 RCTs helped to solve the high heterogeneity and RoB issues found in the previously published article in order to derive a concrete conclusion on the effectiveness of baricitinib in reducing mortality in COVID-19 patients.

## Data Availability

Supplementary Description
The supplementary files are given separately as supplementary file S1 (PRISM flow chart [14] of article inclusion and exclusion).

## Acknowledgement

The author would like to thank the Director General of Health Malaysia for his permission to publish this article, and the Director of the Institute for Medical Research for his support.

## Supplementary Description

The supplementary files are given separately as supplementary file S1 (PRISM flow chart [14] of article inclusion and exclusion).

---

**Supplementary file 1.**
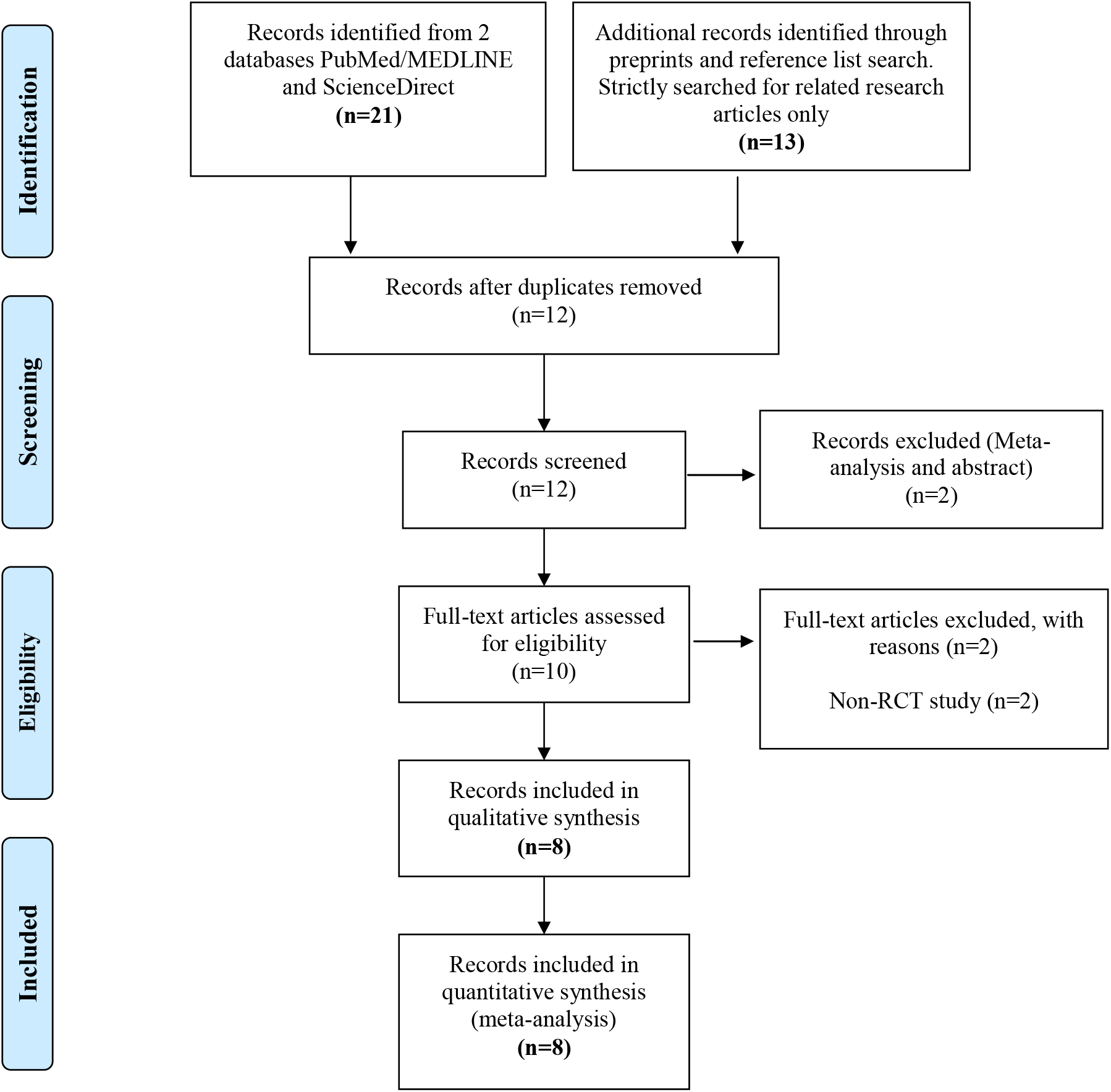
PRISM flow chart [14] of article inclusion and exclusion

## References

[1] Covid-19 coronavirus pandemic. https://www.worldometers.info/coronavirus/

[2] Kalil AC, Stebbing J. Baricitinib: the first immunomodulatory treatment to reduce COVID-19 mortality in a placebo-controlled trial. Lancet Respir Med. 2021; 9: 1349–1351.

[3] Masiá M, Padilla S, García JA, García-Abellán J, Navarro A, Guillén L, et al. Impact of the Addition of Baricitinib to Standard of Care Including Tocilizumab and Corticosteroids on Mortality and Safety in Severe COVID-19. Front Med (Lausanne). 2021; 8: 749657.

[4] Selvaraj V, Finn A, Lal A, Khan MS, Dapaah-Afriyie K, Carino GP. Baricitinib in hospitalised patients with COVID-19: A meta-analysis of randomised controlled trials. EClinicalMedicine. 2022; 49: 101489

[5] Review Manager (RevMan) [Computer program]. Version 5.4, The Cochrane Collaboration, 2020.

[6] GRADEpro GDT: GRADEpro Guideline Development Tool [Software]. McMaster University, 2020

[7] Manoharan S, Ying YL. Does baricitinib reduce mortality and disease progression in SARS-CoV-2 virus infected patients? A systematic review and meta-analysis. Resp Med. 2022; 202: 106986

[8] Manoharan S, Ying YL. Baricitinib for the Management of SARS-CoV-2 Virus Infected Patients. A Systematic Review and Meta-Analysis of Randomised Controlled Trials. Canadian J Infect Dis Med Microb. 2022; Volume 2022: article ID 8332819.

[9] National Institutes of Health (US); Biological Sciences Curriculum Study. NIH Curriculum Supplement Series [Internet]. Bethesda (MD): National Institutes of Health (US); 2007. Understanding Human Genetic Variation. https://www.ncbi.nlm.nih.gov/books/NBK20363/

[10] McLean G, Kamil J, Lee B, Moore P, Schulz TF, Muik A, et al. The Impact of Evolving SARS-CoV-2 Mutations and Variants on COVID-19 Vaccines. mBio. 2022; 13: e0297921.

[11] Open label study. https://www.cancer.gov/publications/dictionaries/cancer-terms/def/open-label-study

[12] FDA Approves Lilly and Incyte’s OLUMIANT® (baricitinib) for the Treatment of Certain Hospitalized Patients with COVID-19. https://investor.lilly.com/news-releases/news-release-details/fda-approves-lilly-and-incytes-olumiantr-baricitinib-treatment.

[13] Sample size table. https://www.research-advisors.com/tools/SampleSize.htm.

[14] Moher D, Liberati A, Tetzlaff J, Altman DG, The PRISMA Group. Preferred Reporting Items for Systematic Reviews and Meta-Analyses: The PRISMA Statement. PLoS Med. 2009; 6: e1000097

[15] Horby PW, Emberson JR, Mafham M, et al. Baricitinib in patients admitted to hospital with COVID-19 (RECOVERY): a randomised, controlled, open-label, platform trial and updated meta-analysis. medRxiv 2022.03.02.22271623

[16] Kalil AC, Patterson TF, Mehta AK, et al. Baricitinib plus Remdesivir for Hospitalized Adults with Covid-19. New Eng J Med. 2021; 384: 795–807.

[17] Marconi VC, Ramanan AV, de Bono S, et al. Efficacy and safety of baricitinib for the treatment of hospitalised adults with COVID-19 (COV-BARRIER): a randomised, double-blind, parallel-group, placebo-controlled phase 3 trial. Lancet Respir Med. 2021; 9:1407–1418

[18] Wesley-Ely E, Ramanan AV, Kartman CE, et al. Efficacy and safety of baricitinib plus standard of care for the treatment of critically ill hospitalised adults with COVID-19 on invasive mechanical ventilation or extracorporeal membrane oxygenation: an exploratory, randomised, placebo-controlled trial. Lancet Respir Med. 2022; S2213-2600(22)00006-6.

[19] Wolfe CR, Tomashek KM, Patterson TF, Gomez CA, Marconi VC, Jain MK, et al. Baricitinib versus dexamethasone for adults hospitalised with COVID-19 (ACTT-4): a randomised, double-blind, double placebo-controlled trial. Lancet Respir Med. 2022: S2213-2600(22)00088-1.

[20] Trøseid M, Arribas JR, Assoumou L, Holten AR, Poissy J, Terzić V, et al., Efficacy and Safety of Baricitinib for the Treatment of Hospitalized Adults with Severe or Critical COVID-19 (Bari-SolidAct): A Randomised, Double-Blind, Parallel-Group, Placebo-Controlled Phase 3 Trial. Available at SSRN: https://ssrn.com/abstract=4172086 or http://dx.doi.org/10.2139/ssrn.4172086

[21] Karampitsakos T, Papaioannou O, Tsiri P, Katsaras M, Katsimpris A, Kalogeropoulos AP, et al. Tocilizumab versus baricitinib in hospitalized patients with severe COVID-19: an open label, randomized controlled trial. Clin Microbiol Infect 2022; S1198-743X(22)00529-8.

[22] Montejano R, de la Calle-Prieto F, Velasco M, Guijarro C, Queiruga-Parada J, Jiménez-González M, et al. Tenofovir Disoproxil Fumarate/Emtricitabine and Baricitinib for Patients at High Risk of Severe COVID-19: The PANCOVID Randomized Clinical Trial. Clin Infect Dis 2022: ciac628.

